# The kidney releases a non-polymerizing form of Uromodulin in the urine and circulation that retains the external hydrophobic patch domain

**DOI:** 10.1101/2021.02.26.21251982

**Authors:** Radmila Micanovic, Kaice A. LaFavers, Kavish R. Patidar, Marwan S. Ghabril, Emma H. Doud, Amber L. Mosley, Angela R. Sabo, Shehnaz Khan, Tarek M. El-Achkar

## Abstract

Uromodulin (Tamm-Horsfall protein, THP) is a glycoprotein uniquely produced in the kidney. It is released by cells of the thick ascending limbs (TAL) apically in the urine, and basolaterally in the renal interstitium and systemic circulation. Processing of mature urinary THP, which polymerizes into supra-molecular filaments, requires cleavage of an external hydrophobic patch (EHP) at the C terminus. However, THP in the circulation is not polymerized, and it remains unclear if non-aggregated forms of THP exist natively in the urine. We propose that an alternative processing path, which retains the EHP domain, can lead to a non-polymerizing form of THP. We generated an antibody that specifically recognizes THP with retained EHP (THP+EHP) and established its presence in the urine in a non-polymerized native state. Proteomic characterization of urinary THP+EHP revealed its C-terminus to end at F617. In the human kidney, THP+EHP was not only detected in TAL cells, but also diffusely in the renal parenchyma. Using immunoprecipitation followed by proteomic sequencing and immunoblotting, we then demonstrated that serum THP has also retained EHP. In a small cohort of patients at risk for acute kidney injury (AKI), admission urinary THP+EHP was significantly lower in patients who subsequently developed AKI during hospitalization. Our findings uncover novel insights into uromodulin biology by establishing the presence of an alternative path for cellular processing, which could explain the release of non-polymerizing THP in the circulation. Larger studies are needed to establish the utility of urinary THP+EHP as a sensitive biomarker of kidney health and susceptibility to injury.

## Introduction

Uromodulin, (also known as Tamm-Horsfall protein, THP) is a glycoprotein uniquely produced by the epithelial cells lining the thick ascending limb (TAL) of the loop of Henle and early distal convoluted tubule (1–3). Its exclusive renal expression, great urinary abundance and phylogenetic conservation point to important physiological roles that are of interest to researchers and clinicians. THP is predominantly sorted to the apex of TAL cells and secreted into the urine as one of the most abundant proteins (4, 5). A lesser, but significant amount of THP is released basolaterally, towards the renal interstitium and systemic circulation (6, 7). The concentration of THP in the circulation is approximately one thousand times lower than the urine (8, 9).

The last two decades have uncovered important roles for urinary and serum THP in regulating renal homeostasis and modulating response and susceptibility to injury. The polymerization of urinary THP is thought to be a key factor in maintaining the impermeability of the TAL segment to water (10). Furthermore, THP filaments play a key role in host defense against ascending infections by binding to bacteria and aiding in pathogen clearance (11, 12). Within the cells where it is produced, THP facilitates the activation of the NKCC2, ROMK and NCC channels, which has key implications on distal handling of salt and hypertension (13–17). Furthermore, THP regulates calcium and magnesium handling by regulating TRPV5 and TRPM6 channels, respectively (18, 19).

THP is also basolaterally released in the renal interstitium and subsequently into the bloodstream (7). In the renal interstitium, THP affects multiple cell types, such as S3 proximal tubules, where it inhibits pro-inflammatory signaling (20, 21), and immune cells, where it regulates the abundance and phagocytic activity of resident mononuclear phagocytes (MPCs) and protects against inflammation following kidney injury (22). THP also has important systemic effects on other organs, such as regulation of granulopoiesis, vascular calcification and mitigation of oxidative stress through inhibition of the TRPM2 channel (23–25).

Many studies have evaluated the role of both serum and urinary THP as biomarkers. A growing body of evidence suggests that the presence of high levels of THP (in the urine or serum) is independently associated with reduced risk of incident acute and chronic kidney disease, kidney disease progression, cardiovascular disease and mortality outcomes (8, 26–34). In addition, THP serum levels are inversely correlated with markers of systemic inflammation (35–37). However, other studies also suggest that urinary THP may be associated with increased risk of hypertension and kidney disease (16). Therefore, understanding the biology and functions of this protein remains incomplete.

Important advancements have been made to delineate the biosynthesis, intracellular maturation along the secretory pathway, excretion and polymerization of mature urinary THP (38–42). In contrast, little is known about the synthesis, cellular sorting and secretion of circulating THP. We have previously demonstrated that in contrast to urinary THP, circulating THP is not aggregated (22). However, it remains unknown how the kidney produces a non-polymerizing form of THP. The THP precursor is comprised of 640 amino acids. The predicted structural domains of THP include four epidermal growth factor (EGF)–like domains, a cysteine rich domain of unknown function (D8C) and a bipartite C-terminal zona pellucida domain (ZP, comprised of ZP-N and ZP-C sub-domains) essential for THP assembly into extracellular urinary polymers of supramolecular structure (2). An extensive hydrophobic interface mediates the ZP-N domain homodimerization, while a structured interdomain linker between ZP-N and ZP-C directs that self-association (38). During intracellular trafficking, THP is kept in a polymerization-incompetent state by the hydrophobic interaction of two motifs, the internal (IHP, residues 430 – 436 and 456 – 462) and external (EHP, residues 598 – 607) hydrophobic patches (43). The IHP is located in the ZP linker region, while the EHP resides between the consensus proteolytic cleavage site (F587) and GPI anchoring site (S614) at the apical membrane on the urinary lumen side. Proteolysis by the type II transmembrane serine protease hepsin at conserved F587 removes the EHP motif, thus permitting correct orientation of ZP-N domains for polymerization to occur in the urine (44, 45). Although monomeric forms of THP in the urine have been previously recovered, these only occurred after treatment with a chaotropic agent and/or as a result of a truncation (22, 46).

Based on our knowledge of the structure of THP and the mechanisms of its polymerization, we aimed to 1) determine how the kidney could natively produce a non-polymerizing form of THP; 2) assess whether this knowledge will advance our understanding of the biology of its apical vs. basolateral release; 3) gauge the potential clinical implications and advantages of measuring such form of THP. We hypothesized that a non-polymerizing form of THP could be present if a molecular event (such as cleavage at an alternative site) causes retention of the EHP and thereby releases a longer form of THP (similar to the precursor protein). Therefore, we designed and optimized tools to detect and measure THP with retained EHP (referred to as THP+EHP) and pursued proteomic sequencing of circulating THP. We also performed preliminary studies to assess the usefulness of measuring THP+EHP in a clinical setting.

## Methods

### Human reference samples

Specimens were obtained from the Biopsy Biobank Cohort of Indiana-BBCI (Indiana University Institutional Review Board approval number: 1906572234). Kidney tissue specimens were from deceased donor nephrectomies. Normal human urine samples were obtained from healthy donors. Normal human serum was purchased from Millipore EMD (# S1-100). Immunoglobulin depleted human serum was purchased from Innovative Research (# IIGGDS100ML).

### Generation of polyclonal anti-peptide antibody for detection of non-polymerizing THP

A rabbit polyclonal antibody recognizing the EHP region of human THP (anti-EHP pAb) was generated by Biomatic (Ontario, Canada). The synthetic peptide corresponding to human THP amino acids 591 – 612 (C-SVIDQSRVLNLGPITRKGVQAT) was used in immunizations (conjugated to keyhole limpet hemocyanin protein) and for antigen-affinity purification of the antibody.

### Western blot

Western blot was performed as described previously (22).The following primary antibodies were used: sheep anti-hTHP (R&D Biosystems #AF5144) pAb and rabbit anti-EHP pAb; secondary antibodies used were rabbit anti-sheep IgG-HRP (Millipore # AP147P) and donkey anti-rabbit IgG-HRP (Millipore #AP182P), respectively, all at starting concentrations of 1 mg/ml. Bands were detected by enhanced chemiluminescence (Pierce Super Signal West Pico kit, #34087) on the ChemiDoc MP imaging system (BioRad, California).

### Blocking with immunizing peptide

Lyophilized immunizing peptide for anti-EHP pAb was dissolved in 10 % DMSO, 90 % PBS to a concentration of 5 mg/ml. It was added to anti-EHP pAb at five times excess by weight and the mixture was incubated with agitation at room temperature for 30 min. The immuno-staining protocol was carried out on two identical samples of human urine (loaded in triplicate) using the non-blocked anti-EHP pAb for one and the blocked antibody for the other as primary antibodies in the Western blot.

### Native PAGE and immunoblot

Native polyacrylamide gel electrophoresis and immunoblot were performed using the Invitrogen 4-16 % NativePAGE Bis -Tris precast mini-gels and protocol, as previously described. (22).

### Urine deglycosylation

Human urine was adjusted to concentrations of RIPA buffer: 25 mM Tris, pH 7.6, 150 mM NaCl, 1 % NP-40, 1% Na-deoxycholate, 0.1% SDS) and treated with protease inhibitors (Pierce Biotechnology #78430). Deglycosylation was performed using Protein Deglycosylation Mix II (New England BioLabs #P6044), according to the manufacturer’s protocol. Reactions were carried out for 1 and 16 hours, respectively, stopped with 4 X LDS sample loading buffer (Life Technologies #B0007) and used in Western Blot analysis.

### Immunoprecipitation of non-polymerizing THP+EHP from the urine

Non-polymerizing THP+EHP was immunoprecipitated from 0.5 L of urine, from which polymerizing THP was previously removed by two consecutive precipitations with 0.58 M NaCl, according to the method of Tamm and Horsfall (47, 48). After removal of salt by dialysis, the urine was concentrated ∼10-fold over Centricon Plus 75 with molecular weight cut off (MWCO) 30 kDa (Millipore #UFC70308) and used in immunoprecipitation. For immunoprecipitation experiments 0.5 mg of rabbit polyclonal anti-THP+EHP antibody was covalently coupled to 50 μl of Protein A agarose beads (Pierce Biotechnologies # 20333) using homobifunctional cross-linking reagent DSS (disuccinimidyl suberate), according to the scaled-up protocol in the Pierce Protein A IgG Plus orientation kit (# 44893).

### Immunoprecipitation of serum THP

THP was immunoprecipitated from 25 ml of commercial immunoglobulin depleted serum treated and adjusted to concentrations in RIPA buffer (25 mM Tris, pH 7.6, 150 mM NaCl, 1 % NP-40, 1% Na-deoxycholate, 0.1% SDS) using either mouse anti-human THP mAb (Santa Cruz # sc-271022) coupled to agarose beads or sheep anti-human THP pAb (R & D Systems # AF5144) followed by Protein A (Pierce Biotechnologies #20333), in 2.5 fold molar excess over the target antigen. The pelleted beads from immunoprecipitation reactions were washed 3 times with RIPA buffer and 3 times with RIPA buffer without detergents (25 mM Tris, pH 7.6, 150 mM NaCl) and submitted for proteomic analysis.

### Proteomic analysis of immunoprecipitated urinary THP+EHP or immunoprecipitated serum THP

Sample preparation, mass spectrometry analysis, bioinformatics, and data evaluation for proteomics experiments were performed in collaboration with the Indiana University Proteomics Core Facility at the Indiana University School of Medicine similarly to previously published protocols (49).

Each immunoprecipitation reaction was treated with 100 ml of 8M urea in 50 mM Tris-HCl, pH 8.5, reduced with 5 mM Tris (2-carboxyethyl) phosphine hydrochloride (TCEP-HCl) for 30 minutes at 37° C and alkylated with 10 mM chloroacetamide (CAA) at room temperature, in the dark, for 30 minutes. The reactions were then diluted with 100 mM Tris pH 8.5, 10 mM CaCl_2_ to 2 M urea and treated with 500 U of PNGase F (NEB P0705L) for 2 hours at 37° C. Proteolytic digestion was accomplished using either trypsin/LysC (Promega #V5072) or chymotrypsin (Roche #11418467001). For the latter digestions the samples were additionally diluted to 0.5 M urea with 100 mM Tris pH 8.0, 10 mM CaCl_2_. The digestions were carried out at 35° C overnight followed by acidification with 0.5% v/v trifluoroacetic acid (TFA). Peptides were then cleaned up with SepPak 18 cartridge (WAT054955, Waters, washed with 1 mL of 0.1% TFA and eluted in 600 µL 70% Acetonitrile 0.1 % formic acid (FA)).

Samples were reconstituted in 20 µl of 0.1 % FA and 2 µl were injected on an Easynano LC1200 coupled with EasySpray 902A column on a Lumos^TM^ Orbitrap mass spectrometer (Thermo Fisher Scientific). Each digestion was run in technical duplicate, once with CID fragmentation and once with EtHCD/HCD fragmentation. In both methods, peptides were eluted on a 160-minute gradient from 6-35 % B, increasing to 85% B over 10 minutes, holding at 85% B for 5 minutes and decreasing to 6% B for 5 minutes (Solvent A: 0.1% FA; Solvent B: 80% ACN, 0.1% FA).

For the CID method instrument parameters were as follows: positive mode, 3 second cycle time with APD and Easy-IC on. Full scan 400-1500 m/z with 120000 resolution, standard AGC and auto max IT, 30% RF lens, 5e3 intensity threshold, charge states 2-7, 60 sec dynamic exclusion. MS2 parameters of 1 m/z quadrupole isolation, 35% fixed CID CE, rapid ion trap detection, 20% AGC and dynamic max IT.

For the EtHCD/HCD method instrument parameters were as follows: positive mode, 3 second cycle time with APD and Easy-IC on. Full scan 400-1600 m/z with 120000 resolution, standard AGC and auto max IT, 30% RF lens, 2e4 intensity threshold, charge states 2-7, 60 sec dynamic exclusion. For MS2 parameters an EtHCD and HCD decision tree was used. For the EtHCD branch charge state and precursor selection ranges were: charge state 3, 400-650 m/z; charge state 4, 400-900 m/z; charge state 5, 400-950 m/z, charge state 6-8; Precursor priority: sort by –highest charge state; lowest m/z priority. MS2 EtHCD parameters were scan priority 1, 1.7 m/z quadrupole isolation, activation type ETD, use calibrated charge dependent parameters and supplemental energy 15% EThcD, orbitrap resolution 50000 standard AGC and auto max IT. For HCD branch: charge state 2; charge state 3, 650-1200 m/z; charge state 4 900-1200 m/z; charge state 5, 950-1200 m/z. MS2 HCD parameters were scan priority 1, 1.7 m/z quadrupole isolation, activation type fixed 30% HCD collision energy, orbitrap resolution 50000 standard AGC and auto max IT.

Raw files were loaded into PEAKS X Pro Studio 10.6 Build 20201221 (Bioinformatics Solutions, (50)) with fragmentation mode, detector and enzyme specified for each file. Precursor ion tolerance was 10 ppm and fragment ion tolerance was 0.6 Da for ion trap data and 0.02 Da for Orbitrap data. Database searches were performed in semispecific mode with the Uniprot_Swissprot Homo sapiens database and common contaminants (20437 entries) with variable modifications of acetylation (K, n-terminal), oxidation (M), phosphorylation (STY), carbamidomethylation (C), and ubiquitination (K). PEAKS PTM and SPIDER searches were enabled to search all de novo peptides above 15% score for over 300 potential PTMs and mutations. A 1 % peptide FDR cutoff (-10lgP≥21.8), PTM Ascore > 10, mutation ion intensity >1 and de novo only score > 80% were applied to the data. In total, 50981 peptides and 1939 protein groups were identified. The mass spectrometry proteomics data (**Supplemental data**) have been deposited to the ProteomeXchange Consortium via the PRIDE partner repository with the dataset identifiers PXD027914 and 10.6019/PXD027914 (51), and a summary has been made publicly available at https://github.com/tashkar/Umod-proteomics-data.

### Purification of human urinary polymerizing THP (pTHP)

Purification of human urinary THP was carried out by salt precipitation of the aggregating THP from the urine, as described by Tamm and Horsfall (47, 48). THP precipitated from urine of normal human donors was subsequently treated with 8 M urea for >12 hours at 4°C with tumbling. It was then concentrated on an Amicon concentrator with 10 kD MWCO (Millipore) and transferred on a Superdex 200 gel filtration column equilibrated with 30 mM phosphate buffer, pH 6.8, in 2 M urea (22). The column was pre-calibrated using specific molecular weight markers (BioRad). THP fractions were collected and the protein concentration in each fraction was determined using a spectrophotometer. Native gel electrophoresis was performed on the recovered peak fractions followed by denaturing and reducing SDS-PAGE. For these experiments, fractions of THP with evidence of polymerization on native gel electrophoresis were pooled to form polymerizing p(THP).

### ELISA for non-polymerizing (THP+EHP) THP detection

Custom sandwich ELISA for non-polymerizing THP detection was developed using a mouse anti-THP monoclonal antibody (Santa Cruz Biotechnology, #sc-271022) as a capture antibody at 1 μg/ml and rabbit anti-EHP polyclonal antibody as the detection antibody at 5 μg/ml. The detection was performed using donkey anti-rabbit IgG–HRP (Millipore EMD AP182P) at 200 ng/ml and 3,3′,5,5′-Tetramethylbenzidine (TMB) Liquid Substrate System as peroxidase substrate (ThermoFisher Scientific, #34028). The assay was performed using Nunc-Immuno^™^MaxiSorp^™^ 96 well solid plates (Sigma-Aldrich, M9410), 5% BSA (ThermoFisher Scientific, #N502) as a blocking solution, PBS/Tween 0.05% /1% BSA as a washing solution, Stop Solution for TMB Substrates (ThermoFisher Scientific, N600) and standard sandwich ELISA protocol. The standard in the ELISA assay was a custom 37 amino acid-long peptide comprised of mouse anti-hTHP mAb epitope (N-terminal) and EHP sequence (C-terminal). Minimum detectable level was between 50 – 100 pg.

### Tissue sectioning, immunofluorescence staining and confocal microscopy

Immunofluorescence staining was performed, as described previously (20, 52) on 50-µm sections of 4% paraformaldehyde–fixed kidneys from a human nephrectomy specimen and sectioned using a vibratome. The following antibodies were used to visualize THP: anti-human uromodulin antibody, antigen affinity-purified polyclonal sheep IgG (R&D Systems #AF5144) and custom made anti-EHP rabbit polyclonal antibody. DAPI was used for staining nuclei and Oregon Green Phalloidin (Life Technologies # O7466) was used for staining F-actin. Confocal microscopy and image acquisition in 3 separate consecutive channels were performed using an Olympus Fluoview confocal microscope system (Japan). For four channel confocal microscopy and image acquisition, a fully automated Leica SP8 confocal with a 20x 0.75 NA Mimm objective was used.

### Quantitation of EHP fluorescence

Quantitation of EHP fluorescence was done on 3 separate fields from a human nephrectomy specimen (20x magnification) using ImageJ. The average intensity (after background subtraction) was measured separately in each tubule (marked as a region of interest) using a brush tool. The background average intensity was measured in negative controls without primary antibody.

### Epitope mapping of mouse anti-THP mAb

Mouse anti-hTHP monoclonal antibody B-2 (Santa Cruz Biotechnology #sc-271022) had a manufacturer’s defined epitope region between amino acids 291 and 425 of human THP. To identify its true epitope a PepScreen approach was used from Sigma Aldrich. A peptide library of 25 peptides, each 15 amino acids in length and with 10 overlapping amino acids, was screened in a direct ELISA using biotinylated mouse anti-hTHP mAb B-2 and streptavidin-HRP as detection. The epitope peptide was identified to be THP amino acids 371-385 and its sequence was used to generate a hybrid standard for the non-polymerizing THP+EHP ELISA.

### ELISA for total THP detection

Total THP was measured in samples of human urine using the Human UMOD ELISA kit from Sigma-Aldrich (#RAB0751).

### Assay for total protein in the urine

Total urinary protein content was determined by modified Lowry protein assay using a detergent compatible micro-assay kit (BioRad # 5000112).

### Patient population and study protocol

The sample population was obtained from a cohort of hospitalized cirrhotic patients who were non-consecutively enrolled into a study evaluating urea metabolism. The study was approved by the Indiana University School of Medicine Institutional Review Board (approval number: 1011004299). The larger study from this cohort investigating the link between admission plasma THP and the risk of acute kidney injury has been previously reported (29). Inclusion criteria for the study included a known diagnosis of cirrhosis and age ≥ 18 years. The diagnosis of cirrhosis was made based on clinical parameters involving laboratory tests, endoscopic or radiologic evidence of cirrhosis, evidence of decompensation (hepatic encephalopathy, ascites, variceal bleeding, jaundice), and liver biopsy, if available. Patients were excluded if they had an unclear diagnosis of cirrhosis, had prior solid organ transplantation, were admitted electively, or if informed consent could not be obtained. We further excluded patients who had active cancer, acute kidney injury (AKI) on admission, hemodialysis at the time of admission, and/or confirmed pregnancy (29). For the purposes of this current pilot study, we identified patients who subsequently developed AKI during hospitalization (defined as a rise in creatinine of 0.3 mg/dL or 50% increase from baseline, as recommended by Kidney Disease Improving Global Outcomes (53)) and had available blood and urine samples stored from the day of admission. 10 patients were identified, and they were matched for age, gender, baseline kidney function, and severity of cirrhosis (Model for End-stage Liver Disease) to another 10 patients who did not develop AKI. The case control matching was finalized *a priori* before any further testing was performed on the plasma or urine. **Table 1** shows the baseline characteristic of this cohort. As expected, there were no significant differences between the two groups for demographics, co-morbid conditions, kidney function (baseline and admission), infections, and severity of cirrhosis.

**Table 1:** Demographics and baseline characteristics of cohort of patients with cirrhosis.

|  | No-AKI | Development of AKI | P-value |
| --- | --- | --- | --- |
|  | N=10 | N=10 |  |
| Age (s.d) | 56.00 (7.21) | 57.00 (9.12) | 0.790 |
| Gender, n (%) male | 6 (60) | 6 (60) | 1.00 |
| Race, n (%) white | 8 (80) | 10 (100) | 0.474 |
| Etiology of Cirrhosis, n (%) |  |  |  |
| Hepatitis C | 2 (20) | 2 (20) | 0.539 |
| Hepatitis C and Alcohol | 2 (20) | 0 (0) |  |
| Alcohol | 2 (20) | 2 (20) |  |
| Non-alcoholic Steatohepatitis | 4 (40) | 5 (50) |  |
| Other | 0 (0) | 1 (10) |  |
| History of Diabetes, n (%) | 4 (40) | 4 (40) | 1.00 |
| History of Hypertension, n (%) | 5 (50) | 4 (40) | 0.470 |
| History of Ascites, n (%) | 6 (60) | 9 (90) | 0.303 |
| Infection on Admission, n (%) | 1 (10) | 4 (40)* | 0.303 |
| Baseline sCr, mg/dL (s.d) | 0.94 (0.30) | 1.00 (0.31) | 0.648 |
| Baseline eGFR, ml/min/1.73m<sup>2</sup>* (s.d) | 84.60 (26.73) | 78.10 (22.27) | 0.562 |
| Admission Heart Rate (s.d) | 85.70 (13.39) | 92.10 (11.46) | 0.266 |
| Admission MAP, mmHg (s.d) | 87.00 (15.50) | 79.20 (10.08) | 0.198 |
| Admission WBC, 10<sup>9</sup>/L (s.d) | 8.10 (4.90) | 8.27 (3.88) | 0.932 |
| Admission Hgb g/dL (s.d) | 10.85 (2.28) | 9.90 (1.47) | 0.284 |
| Admission Platelet Count, k/cumm (s.d) | 106.79 (67.06) | 98.20 (32.46) | 0.363 |
| Admission Serum Sodium, mmol/L (s.d) | 137.10 (3.81) | 131.80 (7.03) | 0.051 |
| Admission sCr, mg/dL (s.d) | 0.98 (0.30) | 1.14 (0.43) | 0.364 |
| Admission eGFR, ml/min/1.73m<sup>2</sup>* (s.d) | 79.53 (26.08) | 70.70 (27.40) | 0.470 |
| Admission Total Bilirubin, mg/dL (s.d) | 3.95 (4.34) | 3.13 (2.93) | 0.627 |
| Admission Albumin, g/dL | 2.95 (0.69) | 2.56 (0.57) | 0.186 |
| Admission INR (s.d) | 1.61 (0.52) | 1.70 (0.63) | 0.729 |
| Admission MELD score (s.d) | 15.50 (6.13) | 17.20 (5.71) | 0.506 |

s.d: standard deviation. sCr: serum creatinine; eGFR: estimated glomerular filtration rate; MAP: mean arterial pressure; MELD: Model for Endstage Liver Disease
\*Based on Chronic Kidney Disease Epidemiology Collaboration creatinine equation

### Statistical analysis

All statistical tests were performed using GraphPad Prism software unless otherwise noted. A two tailed t-test or the Wilcoxon rank sum was used to examine the difference in means for continuous data. A paired t-test was used for samples from the same patient collected at different times. An F-test was performed to test if the variances of the samples were significantly different and a Welch’s t-test with correction was performed if the variances were found to be significantly different. For measurements with more than two groups, an ordinary one-way ANOVA was performed and significance between pairs of two groups was determined using Dunnett’s multiple comparison tests. Only quantitative measurements which fell within the range of the calibrator measurement curve were used for analysis. Outlier measurements were identified according to the Tukey fence method and removed. Chi-squared and Fisher’s exact tests were used to determine differences between categorical variables. Simple linear regressions were used to determine relationships between two continuous variables. Statistical significance was determined at p< 0.05.

## Results

### Establishing the presence of THP+EHP in the urine and its impaired polymerization

The annotated amino acid sequence of full length THP and its structural domains are shown in **Figure 1A-1B**. To test the presence of THP+EHP, we designed, generated, and characterized an anti-peptide antibody that recognizes the EHP sequence (**Figure 2A**). We then investigated and established the presence of THP+EHP in the urine using Western blot (**Figure 2B**). As expected, THP+EHP had a higher molecular weight (>100 kDa) than urinary or purified polymerizing THP (the last two were detected using the sheep anti-THP polyclonal antibody mentioned above). To determine the polymerization state of THP+EHP, we performed native immunoblots, and showed that THP+EHP is present as a dimer in its native state (discrete, single band of molecular weight (MW) around 242 kDa marker), as compared to the multimeric native forms of urinary THP with characteristic ladder pattern (**Figure 2C**).

**Figure 1:**
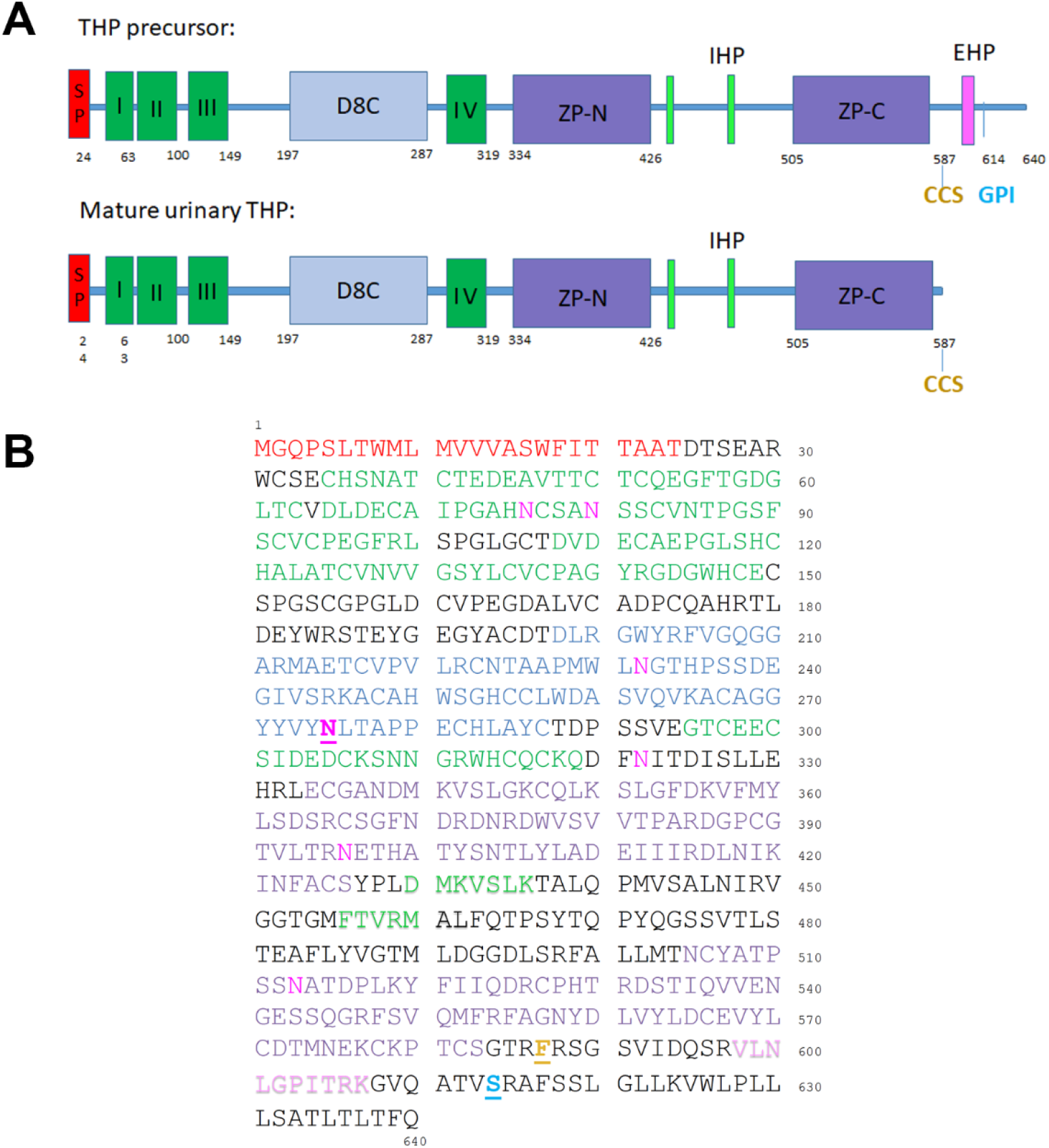
Annotated structure of THP (Uromodulin) and its structural domains. (A)Schematic diagram of THP structure and its functional domains. The top shows initial 640 amino acid THP precursor and the bottom its processed mature urinary form Abbreviations: SP – signal peptide; I, II, III and IV – EGF domains; D8C – cysteine rich domain; ZP – zona pellucida domain; GPI – glycosylphosphatidylinositol anchor at 614; CCS – consensus cleavage site at 587; IHP – internal hydrophobic patch (430 – 436 and 456 – 462); EHP – external hydrophobic patch (598–607). (B) Amino Acid sequence of full-length THP with color coded domain annotation; red – signal peptide; green – EFG-like domains; blue – D8C disulfide bond domain; purple – ZP-N and ZP-C domains; lavender – EHP; light green – IHP; yellow – CCS; cyan –GPI anchor attachment site; magenta – sites of N-linked glycosylation.

**Figure 2:**
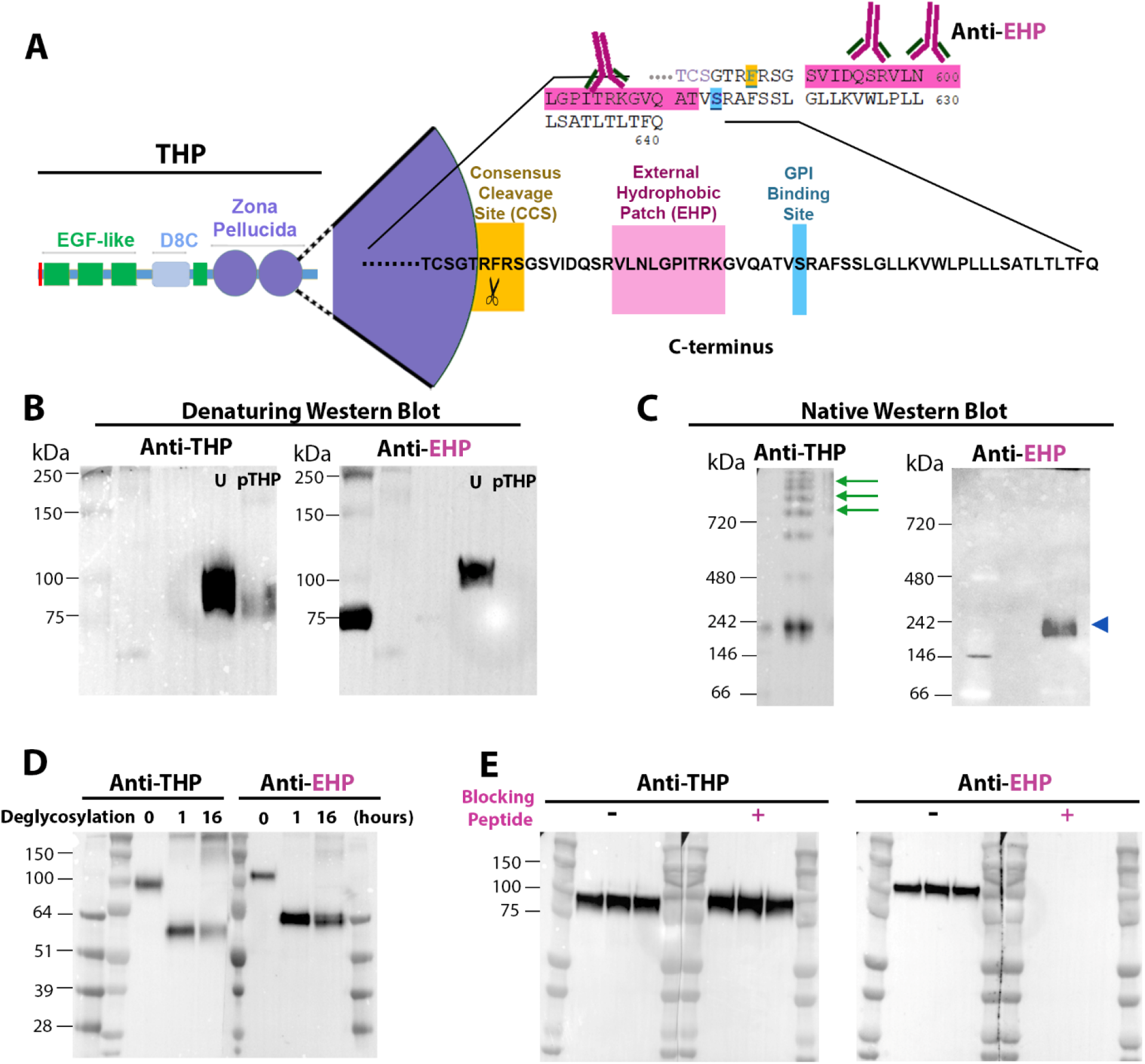
Non-polymerizing THP (Uromodulin) with retained external hydrophobic patch (EHP) is produced in the kidney. (A) Schematic representation of THP (Uromodulin) precursor with annotation of its functional domains, focusing on the C terminus (enlarged). A polyclonal antibody against the indicated sequence encompassing the external hydrophobic patch (EHP) was developed. (B) Left panel shows a denaturing immunoblot where THP with retained EHP (THP+EHP) is detected by anti-EHP in the urine (U). THP+EHP has higher MW than mature urinary THP, the latter detected by re-probing with an antibody (anti-THP) that was raised against mature urinary THP that lacks EHP. No EHP signal is detected in mature urinary polymerizing THP that was purified based on its aggregative properties (pTHP). (C) Native separate immunoblots revealed that THP+EHP is a dimer in its native form (arrowhead, right panel), whereas mature urinary THP without EHP polymerizes into high molecular weight multimers (arrows, left panel). (D) Human urine samples were treated with mixture of deglycosylating enzymes for 0, 1 and 16 hours then resolved on SDS-PAGE followed by immunoblotting. Blots were probed with anti-THP or anti-EHP. (E) Human urine samples were resolved on SDS-PAGE followed by immunoblotting in the presence or absence of EHP blocking peptide.

When the urine was deglycosylated using a mixture of enzymes that remove all *N*-linked, simple *O*-linked glycans, as well as some complex *O*-linked glycans (**Figure 2D**), and probed on the immunoblot, THP+EHP had a MW of ∼65 kDa, whereas the mature urinary secreted THP was of lower MW, ∼ 61 kDa. The latter is in agreement with the predicted MW of 61,518.43 Da calculated based on the primary sequence (T24 – F587). These results support that THP+EHP has a primary sequence that extends beyond the consensus cleavage site at amino acid F587.

The specificity of the anti-EHP antibody (**Figure 2B**) was established by the lack of EHP signal in the purified urinary polymerizing THP. To further test the specificity of the anti-EHP antibody, an immunoblot was performed on urine samples when the probing antibody was blocked with the immunizing EHP containing peptide (**Figure 2E**). As expected, the blocking peptide completely eliminated the signal of THP+EHP, whereas the signal of total THP was unaltered by its presence.

### Proteomic characterization of the C-terminus of urinary THP+EHP

Bottom-up mass spectrometry was carried out on urinary THP+EHP from two different immunoprecipitations using anti-EHP antibody that was covalently attached to Protein A agarose beads. Prior to immunoprecipitations, urine samples were depleted of polymerizing THP by two consecutive NaCl precipitations (**Figure 3A**). The amount of polymerizing THP was diminishing in consecutive salt precipitations whereas the amount of non-polymerizing THP remained constant. We used trypsin digestion in the first proteomic experiment and observed that uromodulin (THP) was by far the most abundant protein with over 400 peptides recovered and >95% coverage (excluding the signal peptide), including the C-terminal region of interest ending with amino acid R615 (**Supplemental data**). In the second proteomic experiment (**Figure 3B**), digestions with both trypsin and chymotrypsin were performed, and over 800 unique uromodulin peptides were identified with > 95% coverage and a final observable amino acid at the C-terminus F617. Cumulatively, these experiments strongly suggest that the C terminus of THP+EHP ends around F617 (**Figure 3C**), which is also in agreement with a molecular weight observed of ∼ 65 kDa in **Figure 2D**, and a predicted MW of 64,750 kDa based on a primary sequence of A23 - F617.

**Figure 3:**
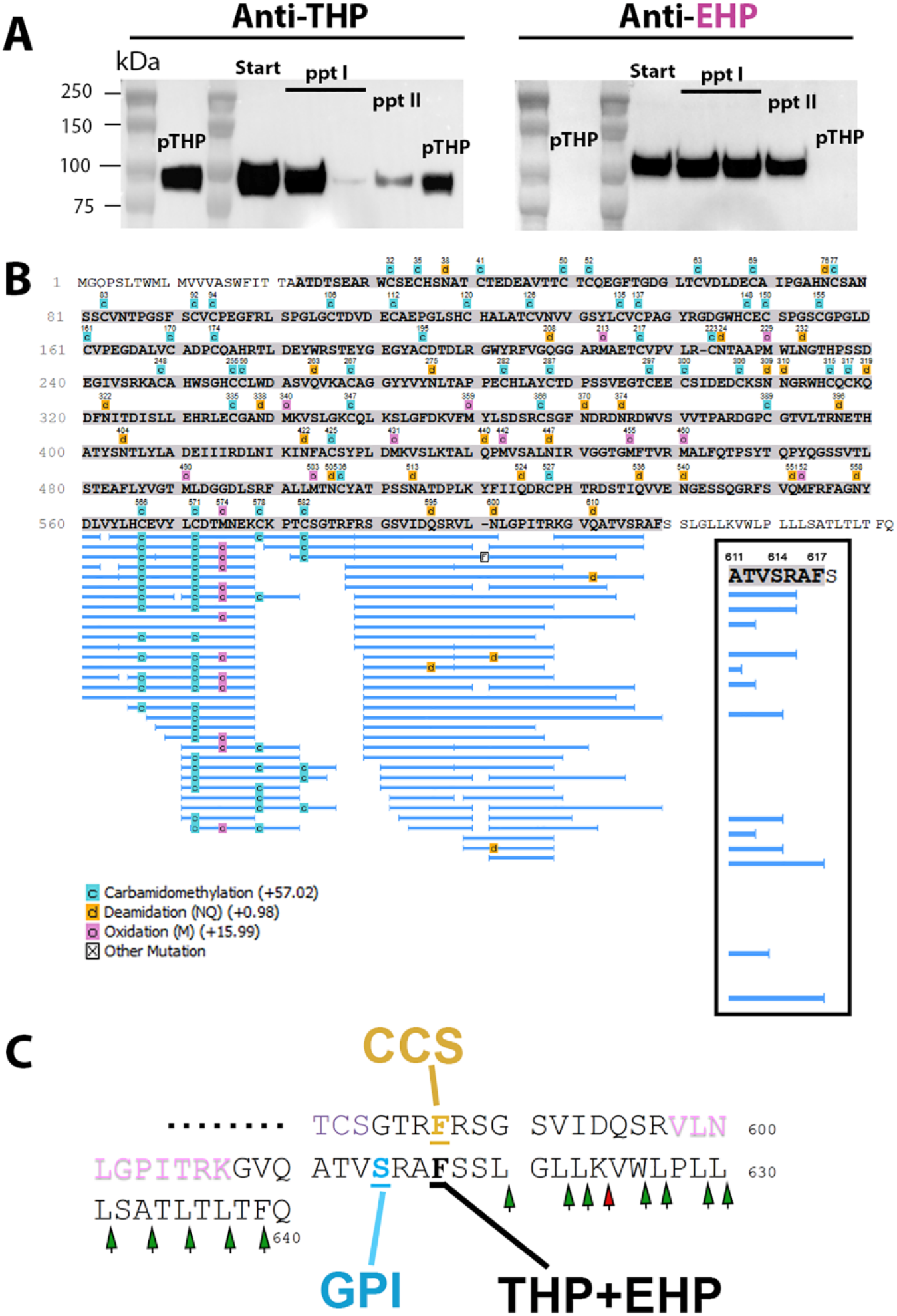
Proteomic characterization of urinary THP+EHP. (A) Polymerizing THP was removed from the urine by two consecutive salt precipitations. The supernatants were resolved on SDS-PAGE followed by Western blotting and probing for THP or EHP. Start - starting urine material; ppt I – urine supernatant after first salt precipitation, drawn immediately (left lane) or 3 days after storage at 4⁰C (right lane); ppt II – urine supernatant after second salt precipitation; pTHP-polymerizing THP that was precipitated. (B) Bottom-up mass spectrometry characterization on immunoprecipitated THP+EHP was carried out with trypsin and chymotrypsin, showing∼ 100% coverage-excluding signal peptide (grey highlight indicates covered sequences) and recovered peptide details are shown for the C-terminus. Enlarged box area focuses on the end coverage of C-terminal region, with the sequence ending at F617 in two chymotryptic peptides. (C) Enlarged peptide sequence of the C terminus, based on (B). Green and red arrows show sites of potential chymotrypsin and trypsin cleavage, respectively. The data searches included de novo peptide identifications and semi-specific protease cleavage which allows for one terminus of an identified peptide to be non-protease specific in order to detect the C-terminal end of THP+EHP with the highest confidence level.

#### Detection of THP+ EHP in the kidney and its localization

We examined the presence of THP+EHP in the human kidney using confocal immunofluorescence and found, as expected, that THP+EHP colocalizes with mature urinary THP in the thick ascending limbs, with a marked expression at the apical domain (**Figure 4A** and **4C**). However, THP+EHP can also be detected more diffusely in the renal parenchyma and in other tubular segments such as proximal tubules (**Figure 4A** and **4D-G**), right lower panel), albeit with a less intense signal.

**Figure 4:**
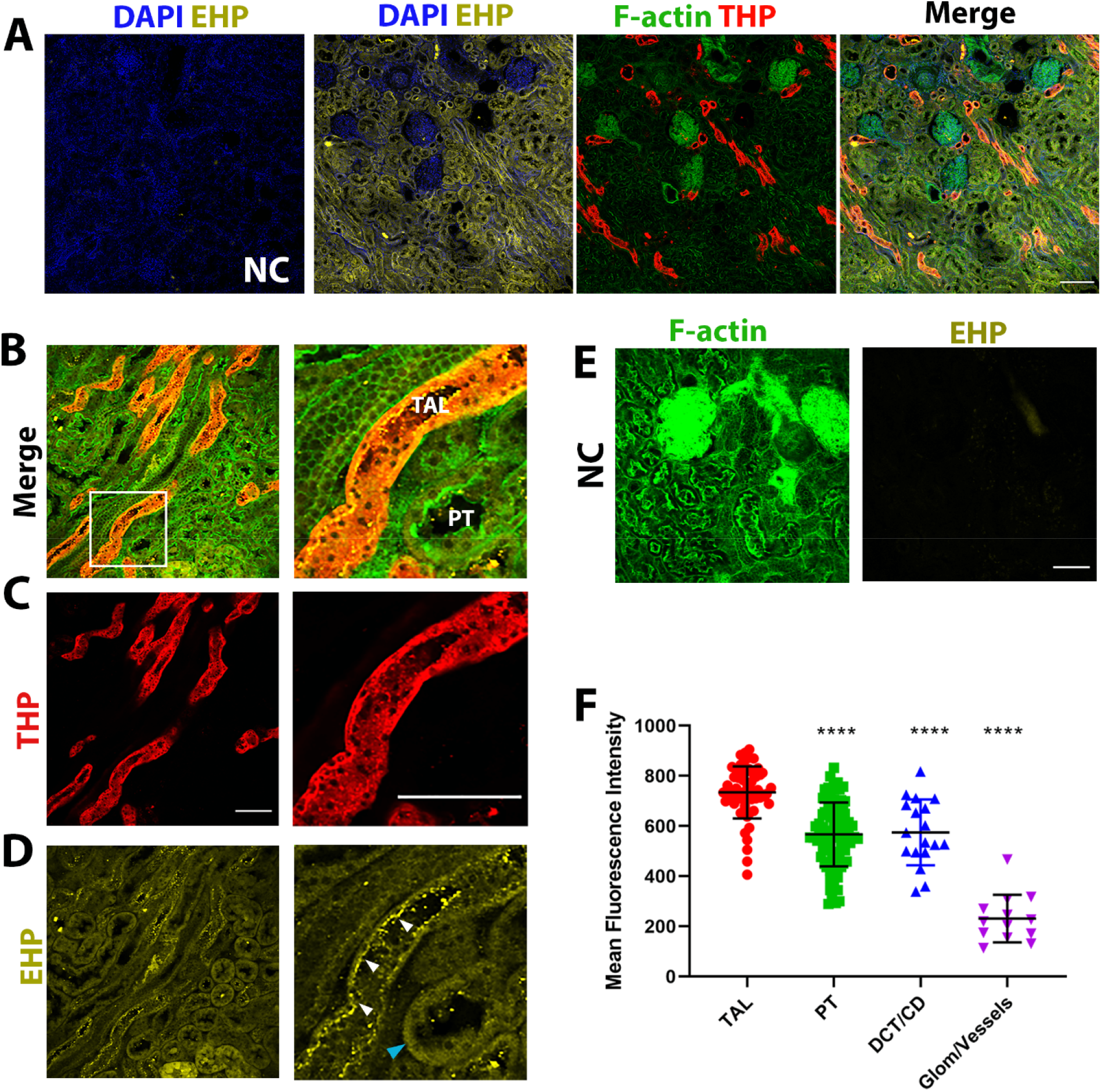
Localization of THP+EHP in the human kidney. (A) Immunofluorescence confocal microscopy of human kidney sections stained to detect THP and THP+EHP (referred to as EHP in the figure). EHP staining appear not only in thick ascending limbs (TAL) but diffusely in other tubules as well, but not in glomeruli. Negative control (NC) for EHP without primary antibody. Scale bar = 200 µm. (B-E) Representative high magnification image from a separate experiment are shown (scale bars = 100µm). The separate channels for THP and EHP from the image in (B) are shown in (C) and (D), respectively. The right panels in (B), (C) and (D) represent a magnification of the area outlined by the box in the left panel in (B). Non-polymerizing, THP+EHP colocalizes with mature THP in TAL tubules, where it is highly expressed at the apical domain (white arrows). However, a significant, albeit less intense THP+EHP signal, can also be detected in other segments such as proximal tubules (blue arrowhead). E shows negative control without primary EHP antibody. (F) Quantitation of THP+ EHP signal per tubule type from 3 separate fields is shown in the graph. Asterisks denote P<0.0001.

#### Detection of EHP in immunoprecipitated serum THP

Immunoprecipitation of immunoglobulin-depleted serum THP was done followed by Western blot analysis using immunoblotting with anti-THP and anti-EHP antibodies (**Figure 5**). Serum THP probed by anti-THP had a comparable MW to serum THP probed with anti-EHP, and also to urinary non-polymerizing THP+EHP. In comparison, total urinary THP (probed with anti-THP) had a broad MW range, which is explained by the detection of both polymerizing THP (with lower MW as shown by the lane with purified polymerizing THP) and non-polymerizing urinary THP+EHP (shown separately when detected by anti-EHP) (**Figure 5**). The specificity anti-EHP is again demonstrated by its inability to detect purified polymerizing THP (pTHP), which lacks the EHP domain. Cumulatively, this data demonstrates that serum THP has a retained EHP domain that is recognized by the anti-EHP antibody.

**Figure 5:**
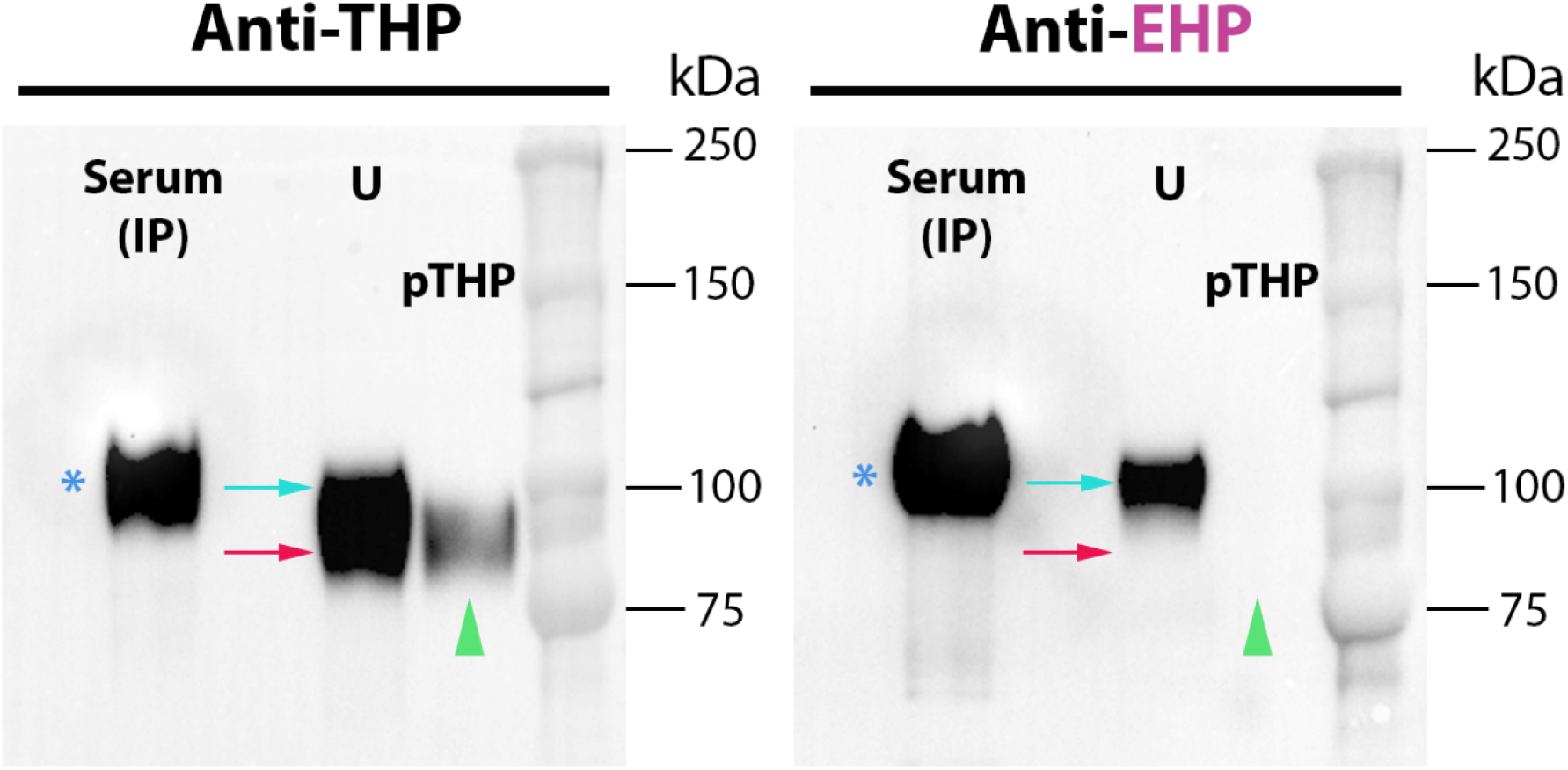
Immunoblotting of immunoprecipitated serum THP demonstrates the presence of EHP. Immunoprecipitation (IP) of serum THP was performed followed by Western blot probed with anti-THP (raised against mature protein, left) or with anti-EHP (right) and compared to urine and purified polymerizing THP (pTHP). Serum THP and THP+EHP have comparable molecular weights to each other (asterisk) and to urinary THP+EHP (cyan arrow). In the urine, anti-THP recognizes total THP, the majority of which is comprised of polymerizing THP (red arrow for the urine band, and green arrowhead for pTHP) and to a lesser extent the non-polymerizing THP+EHP of higher molecular weight (cyan arrow). Anti-EHP recognizes serum THP and only urinary THP+EHP (cyan arrow) and does not recognize pTHP, (green arrowhead) as expected. These findings support the presence of a retained EHP in the serum.

#### Proteomic characterization of the C-terminus of serum THP

To validate the observation that serum THP has a retained EHP, we performed also bottom-up mass spectrometry on serum samples that underwent immunoprecipitation (**Figure 6** and **supplemental data**), similar to what we did for the urine IP experiments. In all the experiments, peptide coverage consistently included the C-terminus (**Figure 6 A -C**) which ended at R606 in two experiments and K607 in a third experiment. These results are consistent with the immunoblotting findings above and prove using an orthogonal approach that serum THP has a retained EHP domain with a C-terminus that ends at amino acid K607, which is also beyond the cleavage consensus site for urinary polymerizing THP at F587 (**Figure 6D**).

**Figure 6:**
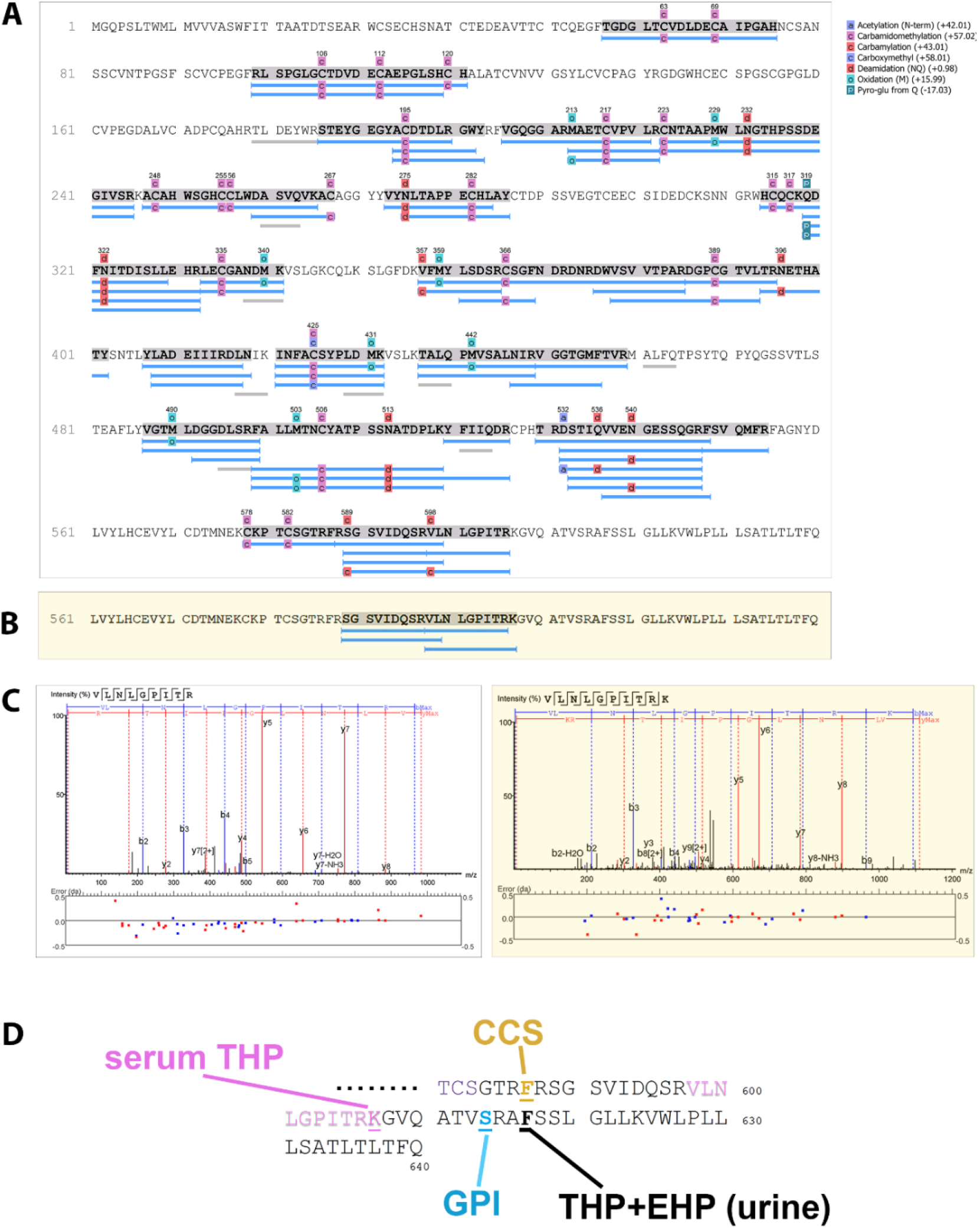
Proteomic characterization of serum THP confirms the presence of EHP. (A) Bottom-up mass spectrometry characterization on immunoprecipitated serum THP was carried out with trypsin and chymotrypsin (grey highlight indicates covered sequences and recovered peptide details are shown). The C-terminal covered region ends at R606. (B) Results from a separate experiment are shown focusing on the end coverage of C-terminal region which ends at K607 (end of EHP domain). (C) MS/MS spectrum view showing fragment matches and relative ion intensities of C-terminal peptides from (A, left) and (B, right) (D) Enlarged peptide sequence of the C terminus, based on (A) and (B), highlighting the recovered C terminal ends of serum THP and urinary THP+EHP.

#### Measuring urinary THP+EHP in patients at risk for AKI

To facilitate the detection and quantitative measurements of THP+EHP, we designed a specific sandwich ELISA. Design and development details are provided in the methods and shown in **Figure 7A**. This effort required the generation of a synthetic hybrid THP+EHP calibrator peptide: <u>DRDNRDWVSVVTPARSVIDQSRVLNLGPITRKGVQAT</u> (**Figure 7A**, right panel). THP+EHP and total THP (assayed using a commercially available ELISA) were then measured in urine samples collected upon hospital admission from a cohort of 20 patients, half of whom subsequently developed hospital acquired AKI. These patients were part of a previously described study of hospitalized patients with liver cirrhosis (29), and they were included *a priori*, based on a case-control design, matching AKI cases with controls with similar age, gender, baseline kidney function, and severity of cirrhosis (**Table 1**). As expected, there were no significant differences between the two groups for demographics, co-morbid conditions, kidney function (baseline and admission), infections and severity of cirrhosis. **Figures 7B** show the measurements in the cohort of urinary total THP and THP+EHP separated by AKI status during hospitalization.

**Figure 7:**
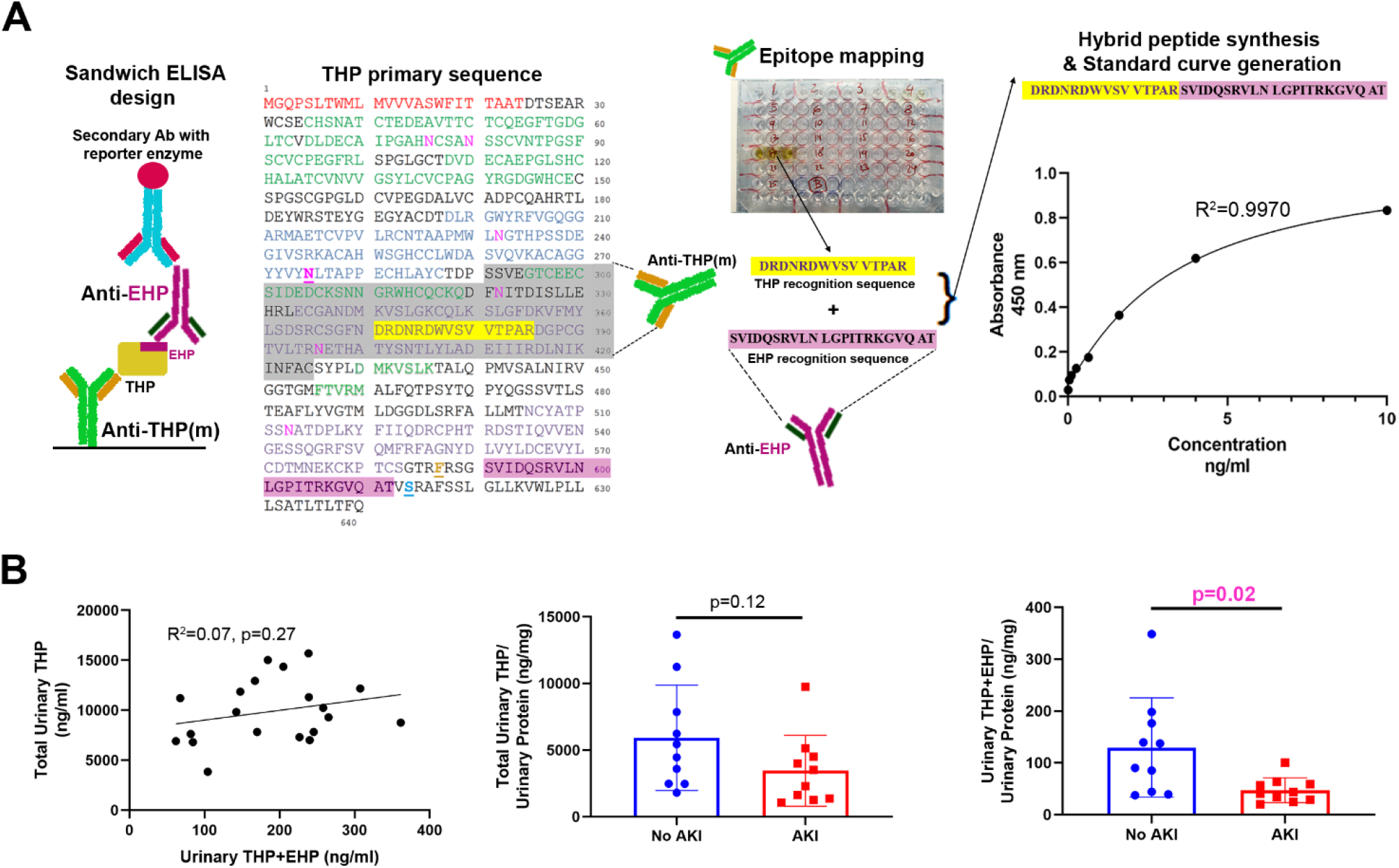
Detection of THP+EHP in the urine of patients at risk for AKI. (A) Design of sandwich ELISA for measurement of THP+EHP. Epitope mapping was performed for a commercially available monoclonal antibody (Anti-THP (m)) and a synthetic hybrid peptide combining the recognizable sequence in mature THP along with EHP sequence was generated for calibration. A representative standard curve using the hybrid peptide is shown in the panel on the right. (B) Levels of non-polymerizing (THP+EHP) and total THP in the urine of a cohort of 20 patients with cirrhosis was measured on admission. Total and non-polymerizing THP did not have a direct significant correlation (left panel). After adjustment to total urinary proteins (there was no statistical difference in total urinary protein between the two groups), only THP+EHP, but not total THP, was lower in patients who subsequently developed AKI (middle and right panels).

##### THP+EHP is significantly decreased in patients who subsequently develop acute kidney injury

There was no significant correlation between THP+EHP and total urinary THP (**Figure 7B,** left panel), suggesting that their expression may be differentially regulated and arguing for an added value in their independent measurements. Specifically, in this small cohort, only non-polymerizing THP+EHP (normalized to total urinary proteins) was significantly decreased on admission in patients who developed subsequent hospital acquired AKI (**Figure 7B,** middle and right panels), suggesting that this form of THP may be a sensitive indicator of kidney health and its susceptibility to acute injury.

## Discussion

In this work, we demonstrate that the kidney produces THP with retained EHP and releases it in the urine and serum. As expected, THP+EHP does not polymerize in its native state, which is consistent with the findings of Schaeffer *et al.* in MDCK cells (43). The presence of THP+EHP at a MW higher than mature urinary THP (slightly higher than 100 kDa when glycosylated and 65 kDa when deglycosylated,) proves that we are not detecting a peptide fragment of THP, but a full-length protein. This was also consistent with our findings in the deglycosylation experiments and the proteomic studies that were performed on immunoprecipitated THP+EHP. In fact, these latter studies demonstrated that the C terminus of this non-polymerizing urinary form extends at least to amino acid F617, which is beyond the GPI linker addition site of S614. Interestingly, hepsin-null mice have been shown to release a non-polymerizing isoform of THP that ends at K616 (using trypsin or Asp-N digestion) (44), which matches the homologous R615 position obtained in our experiments when we only used trypsin (not shown in main figures but included in **Supplemental data**). Therefore, in agreement with these findings in mice (44, 54), our studies support that the release of THP+EHP in humans is not non-specific, but likely occurs though an alternative pathway that is independent of GPI anchoring and cleavage by hepsin.

We propose that this alternative pathway of releasing non-polymerizing THP occurs intracellularly, whereby the carboxy-terminus is proteolyzed without addition of the GPI anchor. Such a mechanism has been described for other proteins with both membrane (GPI-anchored) and soluble forms such as CD14 and folate receptor type β (55, 56). It has been suggested that the divergence at the carboxy-terminal signal for GPI modification may provide an adaptive advantage when there is a need for the functional protein both at the cell surface and in the extracellular environment (56). This could apply to THP, particularly because the functions of its polymerized form (dependent on GPI anchoring and cleavage by hepsin for polymerization (12, 41, 42, 44)) are distinct from non-polymerized form (immunomodulatory and effects in the circulation (1, 22, 24)). THP+EHP levels do not correlate with total THP, suggesting that their production may be independently regulated. In addition, there is emerging evidence supporting that circulating THP (which is non-polymerized) has distinct clinical correlates compared to total urinary polymerized THP, suggesting differential regulation (57). Further studies are needed to elucidate the cellular mechanism underlying the release of THP+EHP, and the analytical tools generated by our work will be useful in performing these studies. We note that the release of non-polymerizing THP+EHP remains a minor path for urinary secretion, as most of the THP present is in the form of mature THP (without EHP) destined for polymerization.

In the human kidney, we demonstrated the colocalization of THP+EHP with mature THP in TAL cells. The marked apical localization suggests that there is still significant apical sorting despite the presence of an alternative processing pathway that may be GPI independent. The detection of THP+EHP in other tubules suggests a possible basolateral release and uptake by other tubular segments within the kidney. Indeed, we previously showed using immuno-electron microscopy that THP could be detected in the interstitium and at the basolateral aspect of proximal tubules neighboring to TAL (7). It is possible that retaining the EHP in this alternative processing pathway could be an important mechanism by which THP is released into the kidney interstitium, parenchyma and circulation. If this proposition is true, we would then expect a significant detection of THP+EHP in the circulation.

Indeed, immunoprecipitated serum THP was detected using both anti-THP and anti-EHP antibodies around the same molecular weight as urinary THP+EHP. This supports that a major portion of circulating THP has retained EHP. To confirm this, we report the first proteomic sequencing of the C terminus of serum THP, which was performed in three separate experiments (two were shown in **Figure 6**). We successfully demonstrated that the serum THP retains the EHP and ends most likely at amino acid K607. The fact that we detected THP using the anti-THP and anti-EHP antibodies in the serum at a comparable MW to that of urinary THP+EHP cross-validates the proteomics findings in that we are not detecting a proteolytic fragment of THP in the serum.

We propose that TAL cells use the same hepsin-independent alternative processing path for releasing both urinary THP+EHP and basolateral/circulating THP. The recovered primary sequence of serum THP is slightly shorter than urinary THP+EHP by proteomic characterization (K607 vs. F617, this small difference is unlikely to be detected by MW shifting on a western blot since it translates to ∼1kDa, assuming the glycosylation is the same). This observation could suggest additional cleavage in the kidney or systemically. Indeed, such molecular path is reminiscent to processing of hormones, that are found within a larger protein (a preprohormone) and whose production usually follows specific rules (58, 59). Our data suggest a plausible model, whereby serum THP is processed at double basic residues R606-K607, that could serve as a recognition site for endoproteinase (58, 59). The latter is a trypsin-like, serine protease processing enzyme, that could cleave the precursor THP to liberate circulating THP. However, we cannot rule out a different processing path for serum THP that still involves a GPI anchoring at S614 and further cleavage at K607. In addition, we cannot completely rule out that we were not able to recover a peptide between K607 and F617 in the serum. We also cannot exclude that differences in glycosylation could occur between basolateral vs. apically released THP+EHP, which could cause slight shifts in MW. We believe that these findings will be foundational for future studies that will further elucidate the molecular mechanisms of the basolateral release of THP. The data presented on serum THP also indicates that serum and urinary THP are likely independently regulated, as discussed earlier. Furthermore, because of the mounting interest in serum THP and its role in kidney and cardiovascular disease, understanding these fundamentals about its release will be key to further elucidate the functions of this protein and the potential of altering its levels for therapeutic purposes.

In its native form, urinary THP+EHP forms a dimer (**Figure 2C**). This finding is consistent with our previous work where we isolated a truncated form of urinary THP lacking part of the ZP domain (absent ZP-C domain and ending around amino acid 434) and showed that it could dimerize (22). It is possible that dimerization could be dependent on the ZP-N domain, independently from the ZP-C domain (60, 61). Dimerization could also be dependent on other factors, since it has been reported with other proteins harboring EGF-like domains (62). Further studies are needed to elucidate the mechanism of dimerization, and whether it is of biological relevance.

Our findings may be relevant for future clinical applications by measuring non-polymerizing THP as a biomarker for nephron function, reserve, and risk assessment. The data from the small clinical cohort suggest that THP+EHP may offer some advantages over total THP for risk assessment of kidney injury. Being non-polymerized, the levels of THP+EHP may be less susceptible to fluctuation that could be introduced by changes in the polymerization state. The ELISA developed in this study (**Figure 7A**) is less likely to detect proteolytic fragments, because of the requirement by this assay for an intact sequence between the epitopes used for capture and detection.

Furthermore, the binding of antibodies may be more consistent in the setting of a non-aggregated protein, thereby causing less variability in the readout, and improving the accuracy and robustness of the measurements. Therefore, we propose that measurements of THP+EHP may offer improvements in representing functional nephron mass, reserve and the changes induced by disease, compared to total THP. Further studies are needed to test this hypothesis and establish the susceptibility of THP+EHP to degradation compared to mature urinary THP (63). We also like to caution against comparing the absolute levels of THP+EHP obtained by this ELISA to total THP obtained by commercial ELISA, because a synthetic peptide was used for calibration in the former, as compared to a protein in the latter. Therefore, we cannot exclude that differences in conformation between a peptide and a full-length protein will differentially affect the ability to bind an antibody. However, we do not expect that this will alter the relationships derived individually by each ELISA. We are also in the process for optimizing an ELISA for THP+EHP in the serum, but this may not necessarily add any advantages since we showed that serum THP has retained EHP, so measurements of total THP in the serum may be a reasonable surrogate for serum THP+EHP.

It is possible that our assay could be cross-reacting with Glycoprotein2 (GP2), a protein with 45% sequence identity to THP, including the EHP region. In fact, GP2 peptides have been detected a very low abundance in human urine (64). However, it is very unlikely that we are detecting GP2 by our assay since the molecular weight of GP2 is ∼ 75-80 kDa (65) and the EHP antibody is recognizing a 100 kDa protein under denaturing conditions both in the urine and the serum. Furthermore, in the proteomic characterization of the IP samples, GP2 peptides in the urine were extremely low (8 total peptides, 5 unique peptides with 6% coverage) and not enriched compared to what was previously described in the urine (1-14 peptides with 2-26% coverage) (64). In the IP samples from the serum, no GP2 unique peptide was detected in any of our experiments, thereby nearly excluding the possibility that the EHP signal in the serum is related to cross-reactivity with circulating GP2.

Our study is limited by the small sample size and requires independent validation in a larger study. We are underpowered to detect other expected outcomes and acknowledge that this is only pilot data. We used this particular small cohort because it was well characterized clinically (29) with appropriate sample storage and accurate in-hospital follow up, which gave us the opportunity to study the risk of subsequent AKI for patients whose lab test were performed on admission. This cohort also gave us the opportunity to perform a case-control design and generate this proof-of-principle data that is needed as a rationale for a larger study. Although our groups were well matched for comorbidities and severity of liver disease, the serum sodium was trending lower in the AKI group. Serum sodium is associated with increased mortality in cirrhosis (66), but the value of admission sodium in predicting hospital acquired AKI in patients with cirrhosis is not established (67). The value of measuring THP+EHP as a sensitive and specific predictor for risk of AKI needs to be performed on a large scale, and for the time being, it is premature to advocate its use outside of clinical research settings.

In conclusion, we provide novel evidence that the kidney produces a non-polymerizing form of uromodulin (THP) by retaining the EHP domain and releasing it in the urine and serum. We show for the first time that circulating THP has an EHP domain, which explains its non-polymerizing properties. Our findings suggest that this form of THP may be independently regulated and provide new insights into the biology of its synthesis and release, which will spur further research. We also provide proof-of-concept data that measurements of this non-polymerizing form could provide an independent value in assessing the risk of kidney injury, a finding that requires validation in larger studies.

## AUTHOR CONTRIBUTIONS

RM: designed reagents, designed and performed experiments, analyzed data and drafted manuscript

KL: designed and performed experiments, analyzed data and drafted manuscript

KP: designed and performed clinical studies, collected specimens

MSG: designed and performed clinical studies, collected specimens

EHD: performed proteomics experiments and data analysis

ALM: performed proteomics experiments and data analysis

ARS: performed imaging experiments

SK: performed tissue processing and staining

T M E: oversaw and designed entire study, performed and analyzed experiments and drafted manuscript.

All authors: edited and approved final draft.

## Data Availability

All the data has been presented in the manuscript and methods

## ACKNOWLEDGEMENTS

This work was supported by funding from the Veteran Affairs (Merit Award to T.M.E.), the National Institute of Diabetes and Digestive and Kidney Diseases (1R01DK111651 to T.M.E), and P30DK079312-Indiana University O’Brien Center for Advanced Renal Microscopic Analysis. The authors would like to thank Drs. Katherine J. Kelly and Daria Barwinska for assistance in experiments.

The mass spectrometry work performed in this work was done by the Indiana University School of Medicine Proteomics Core. Acquisition of the IUSM Proteomics core instrumentation used for this project was provided by the Indiana University Precision Health Initiative. The proteomics work was supported, in part, by the Indiana Clinical and Translational Sciences Institute, which is funded, in part, by Award Number UL1TR002529 from the National Institutes of Health, National Center for Advancing Translational Sciences, Clinical and Translational Sciences Award and the Cancer Center Support Grant for the IU Simon Comprehensive Cancer Center (Award Number P30CA082709) from the National Cancer Institute."

